# The SARS-CoV-2 antibody landscape is lower in magnitude for structural proteins, diversified for accessory proteins and stable long-term in children

**DOI:** 10.1101/2021.01.03.21249180

**Authors:** Asmaa Hachim, Haogao Gu, Otared Kavian, Mike YW Kwan, Wai-hung Chan, Yat Sun Yau, Susan S Chiu, Owen TY Tsang, David SC Hui, Fionn Ma, Eric HY Lau, Samuel MS Cheng, Leo LM Poon, JS Malik Peiris, Sophie A Valkenburg, Niloufar Kavian

## Abstract

**Background:** Children are less clinically affected by SARS-CoV-2 infection than adults with the majority of cases being mild or asymptomatic and the differences in infection outcomes are poorly understood. The kinetics, magnitude and landscape of the antibody response may impact the clinical severity and serological diagnosis of COVID-19. Thus, a comprehensive investigation of the antibody landscape in children and adults is needed.

**Methods:** We tested 254 plasma from 122 children with symptomatic and asymptomatic SARS-CoV-2 infections in Hong Kong up to 206 days post symptom onset, including 146 longitudinal samples from 58 children. Adult COVID-19 patients and pre-pandemic controls were included for comparison. We assessed antibodies to a 14-wide panel of SARS-CoV-2 structural and accessory proteins by Luciferase Immunoprecipitation System (LIPS).

**Findings:** Children have lower levels of Spike and Nucleocapsid antibodies than adults, and their cumulative humoral response is more expanded to accessory proteins (NSP1 and Open Reading Frames (ORFs)). Sensitive serology using the three N, ORF3b, ORF8 antibodies can discriminate COVID-19 in children. Principal component analysis revealed distinct serological signatures in children and the highest contribution to variance were responses to non-structural proteins ORF3b, NSP1, ORF7a and ORF8. Longitudinal sampling revealed maintenance or increase of antibodies for at least 6 months, except for ORF7b antibodies which showed decline. It was interesting to note that children have higher antibody responses towards known IFN antagonists: ORF3b, ORF6 and ORF7a. The diversified SARS-CoV-2 antibody response in children may be an important factor in driving control of SARS-CoV-2 infection.

## Introduction

The SARS-CoV-2 virus emerged in December 2019 and given the lack of pre-existing immunity has caused a pandemic. The spectrum of COVID-19 disease ranges from asymptomatic to lethal infection. It is now evident that the immune response plays a major role in the pathogenicity and outcome COVID-19^1^. Children are minimally affected clinically by SARS-CoV-2 and the morbidity and mortality observed in adults increases progressively with age, although the viral loads in the respiratory tract are reportedly comparable between children of all ages and adults^2^. The multisystem inflammatory syndrome (MIS-C) that appears in children after infection with SARS-CoV-2 is a rare exception (0.002% of cases) to the generally milder clinical disease observed^3^. Symptoms such as fever, cough, pneumonia and elevated C-reactive protein which are associated with disease severity, are less common in children^4^. The majority of children are asymptomatic and only a minority develop mild symptoms (most commonly fever, cough, pharyngitis, gastrointestinal symptoms and anosmia)^4^, creating difficulties in identifying pediatric cases and in contact tracing. These observations are in contrast with other respiratory virus infections (respiratory syncytial virus (RSV), influenza virus) where children are affected more commonly and more severely compared to adults^5^. Recently, a small family case study by Tosif *et al*., indicated that children can mount an immune response with detectable antibodies to SARS-CoV-2 without preceding detectable viral load, therefore avoiding the development of a symptomatic SARS-CoV-2 infection^6^. The clinical differences observed in children and adults upon SARS-CoV-2 infection may be explained by several immune factors (amongst other clinical or physiological factors), such as pre-existing and cross-reactive immunity to common cold coronaviruses (CCC)^7^ with more recent exposure likely in children, immuno-senescence and inflammatory state^8^, innate immune responses, presence of auto-antibodies^9^, and “trained immunity” as a result of off-target effects of live attenuated vaccines for other infections^5,10^.

Although a growing number of SARS-CoV-2 serology tests are currently in use worldwide and are the basis of the SARS-CoV-2 infection rate data, there is an absence of information on serological responses in children with RT-PCR confirmed SARS-CoV-2 infection. Large epidemiological studies report that children only represent 1-2% of all SARS-CoV-2 cases^11,12^ but this may be and underestimate because of differences in the development of the antibody responses to SARS-CoV-2 in children and pandemic response measures such as school closures. Serology is crucial for determining infection attack rates in the population and for assessing the response to a future vaccine to curb the global pandemic. Most serological tests available rely either on neutralizing antibodies or on the detection of antibodies targeting the Spike (S) or the Nucleocapsid (N) proteins of the virus^13^. We have previously demonstrated that antibodies that are directed against non-structural proteins of the virus, namely ORF3b and ORF8, can be used for accurate diagnosis of SARS-CoV-2 infection^14^. Whilst the cellular immune profile of children appears comparable to adults in a small case study^6^, there are no reports on the humoral antibody landscape and kinetics in pediatric cases. There is a lack of information on the SARS-CoV-2 antibody responses in adults or children to the virus accessory proteins, for instance ORF3b, ORF6 and ORF7a, which have been reported to be potent interferon antagonists that may play a role in immune evasion^15-17^. A finely tuned and balanced antibody response may impact COVID-19 outcomes, and the breadth and magnitude of the landscapes of antibody responses to non-structural proteins may indicate the extent of virus replication and thus immune control.

In the present study, children and adults with SARS-CoV-2 RT-PCR confirmed infection were used to study the antibody landscape to a comprehensive panel of 14 different structural and accessory proteins by LIPS. We tested a population of 122 infected children and 36 infected adults in Hong Kong, including 58 patients with longitudinal samples (2 to 4 time points, with a range 0 to 206 days post infection) to determine the longevity of the antibody responses. Furthermore, due to intensive contact tracing and case-finding measures in Hong Kong, asymptomatic cases with RT-PCR confirmed infections have been identified and their antibody responses are also profiled. These samples were collected between April to November 2020 and include predominantly the third wave of infection cases in Hong Kong corresponding to the period of July to September 2020. Our data reports to date, the most extensive data on the landscape and kinetics of antibody responses in COVID-19 children.

## Results

### SARS-CoV-2 infected children have lower levels of antibodies than adults to all structural proteins, except E

We used the unbiased and quantitative LIPS platform to determine the antibody titres to an extensive panel of 14 antigens from structural and non-structural SARS-CoV-2 proteins in plasma samples from a cohort of infected children, in comparison to adults and controls in Hong Kong (Table 1).

**Table 1.** Subject cohorts details

|  |  | *Pediatric COVID-19* |  | *Adults COVID-19* |  | *Negative* |  |
| --- | --- | --- | --- | --- | --- | --- | --- |
|  |  | N (%) |  | N (%) |  | N (%) |  |
| *Patients* | *Samples* | 254 |  | 36 |  | 33 |  |
|  | *Individuals* | 122 |  | 36 |  | 33 |  |
| *Symptoms* | *Asymptomatic* | 44 (36%) |  | 9 (25%) |  | - |  |
|  | *Mild/Severe* | 78/0 |  | 25/1 |  | - |  |
| *Age* |  | Mean±stdev | 10.8 ± 4.9<br>years | Mean±stdev | 34 ± 17.1<br>years | Mean±stdev | 29.3 ± 16.3<br>years |
|  |  | 0-8 years | 45 (37%) | Median | 31 | Adults<br>(N=13) | 30 - 65 |
|  |  | 8-12 years | 33 (27%) | Min | 18 | Pediatrics<br>(N=20) | 11 - 18 |
|  |  | 12-18 years | 44 (36%) | Max | 71 |  |  |
| *Gender* | *Female* | 49 (40%) |  | 10 (28%) |  | 17 (51%) |  |
|  | *Male* | 73 (60%) |  | 26 (72%) |  | 16 (49%) |  |

Our first data set represents COVID-19 cases of mixed timepoints and symptoms to determine the overall antibody landscapes in children (mean+/-stdev: 39±47 days, range: 0-206 days), adults (mean±stdev: 54±20 days, range: 24-123 days) and negative controls. S and N antibodies are the most widely used antibodies in COVID-19 serology testing worldwide. We therefore first determined the levels of antibodies to different S sub-units by using 3 different S constructs in the LIPS assay: S1 which contains the RBD domain, S2 and the S2’ cleaved subunit (Figure 1). The levels of the two Spike antibodies, S1 and S2’ were markedly lower in children compared to the adult cohort (p<0.0001, p=0.0015 and p<0.0001 respectively, Figure 1ac), whereas no difference was observed for S2 antibodies (Figure 1b). Moreover, N antibodies were significantly elevated in the pediatric COVID-19 cohort relative to negative controls (2.45e5±2.8e5 LU versus 4.15e4±1.5e4 LU (p=0.0045), but did not reach the levels observed in the COVID-19 adult cohort that were almost half a log above (5.45e5±3.0e5 LU adult COVID-19 cohorts, p<0.0001, Figure 1d).

**Figure 1.**
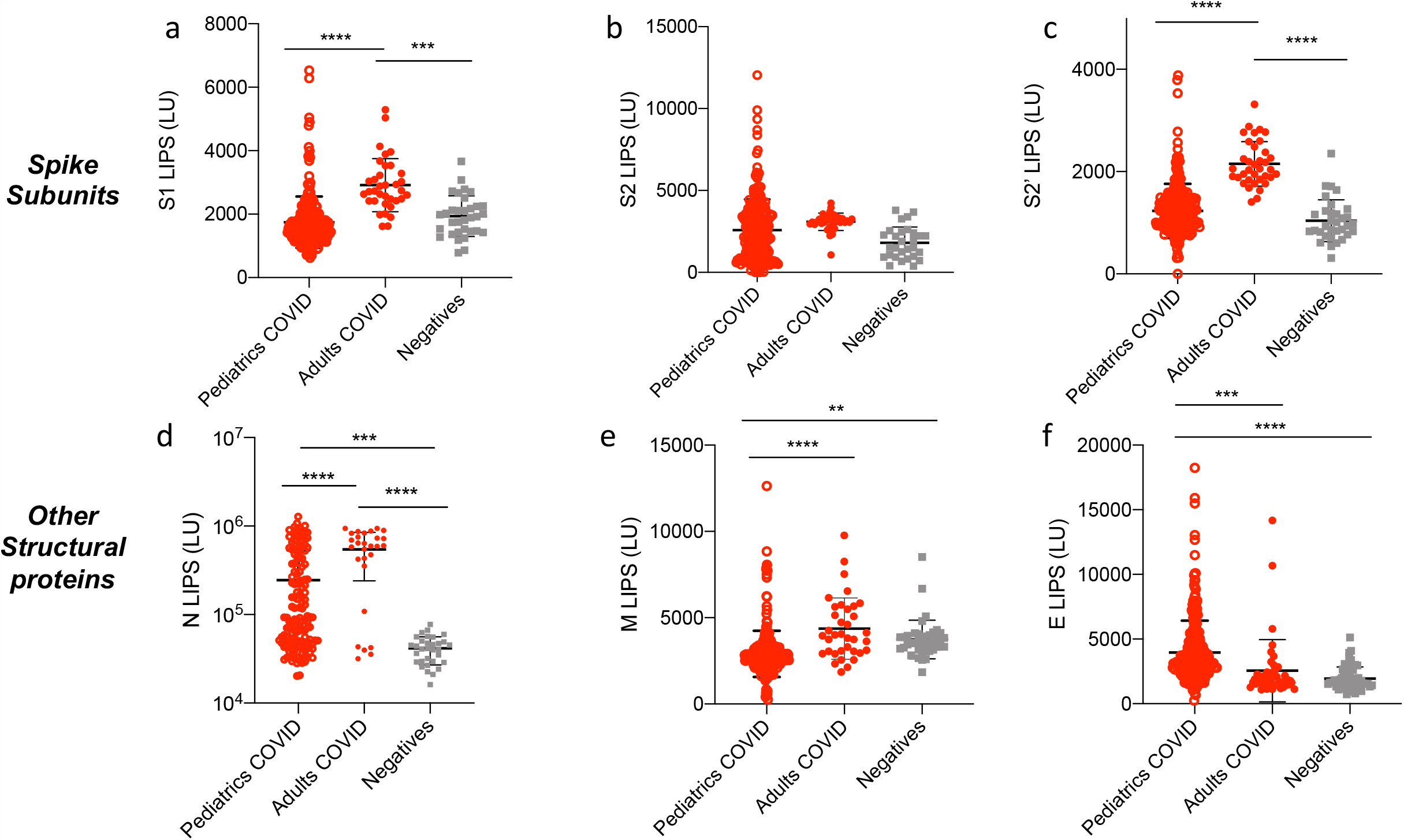
Comparison of antibody responses to SARS-CoV-2 structural proteins in children and in adults with COVID-19. Antibodies against the SARS-CoV-2 structural proteins Spike S1 subunit (S1) (a), Spike S2 subunit (b), Spike S2’ subunit (c), Nucleocapsid (N) (d), Membrane (M) (e), and Envelope (E) (f) were measured by LIPS from samples from pediatrics COVID-19 (n=254) or adult patients (n=36), and negative controls (n=33). Background no plasma values were subtracted. Experiments were repeated twice. All data represents individual responses, and the mean +/- stdev. Two-sided P values were calculated using the Mann-Whitney U test. * shows statistical significance between COVID-19 patients versus negative controls. **p<0.01, ***p<0,001, **** p<0,0001.

We also assessed by LIPS antibodies to other structural proteins Matrix (M) and Envelope (E), which are not widely measured in serology. As for S1, S2’, and N, we found that M antibody levels were lower in the COVID-19 children compared to the adult COVID cohort (p<0.0001, Figure 1e) but were significantly higher that seen in controls. E antibodies followed an inversed trend as they were significantly elevated in the pediatric COVID-19 cohort (Figure 1f) compared to both adult COVID-19 (p=0.0006) and negative controls (p<0.0001).

### Increased breadth of accessory antigen targets in the pediatric COVID-19 population

We next investigated the levels of antibodies directed against the non-structural protein 1 (NSP1) and all the ORFs proteins of the virus. In line with our previous study^14^, adults with COVID-19 displayed elevated levels of NSP1, ORF3a, ORF3b, ORF7a, ORF7b, and ORF8 antibodies compared to negative controls (p<0.0001, p<0.0001, p<0.0001, p=0.05, p=0.0009, p<0.0001, Figure 2a-c and e-g). Again, no detectable levels of ORF6 and ORF10 antibodies were detected in the adult COVID-19 population (p=0.8691 and p=0.999 respectively, Figure 2d and 2h). We observed that the COVID-19 children cohort displayed significantly lower levels of ORF3a, ORF7a, ORF7b antibodies than the adult COVID-19 cohort (p=0.0001, p<0.0001 and p<0.0001 respectively, Figure 2b, e-f). The magnitude of antibody responses to NSP1, ORF3b and ORF8 were comparable in the pediatric COVID-19 and adult COVID-19 populations, and significantly elevated compared to negative controls (Figure 2c and g).

**Figure 2.**
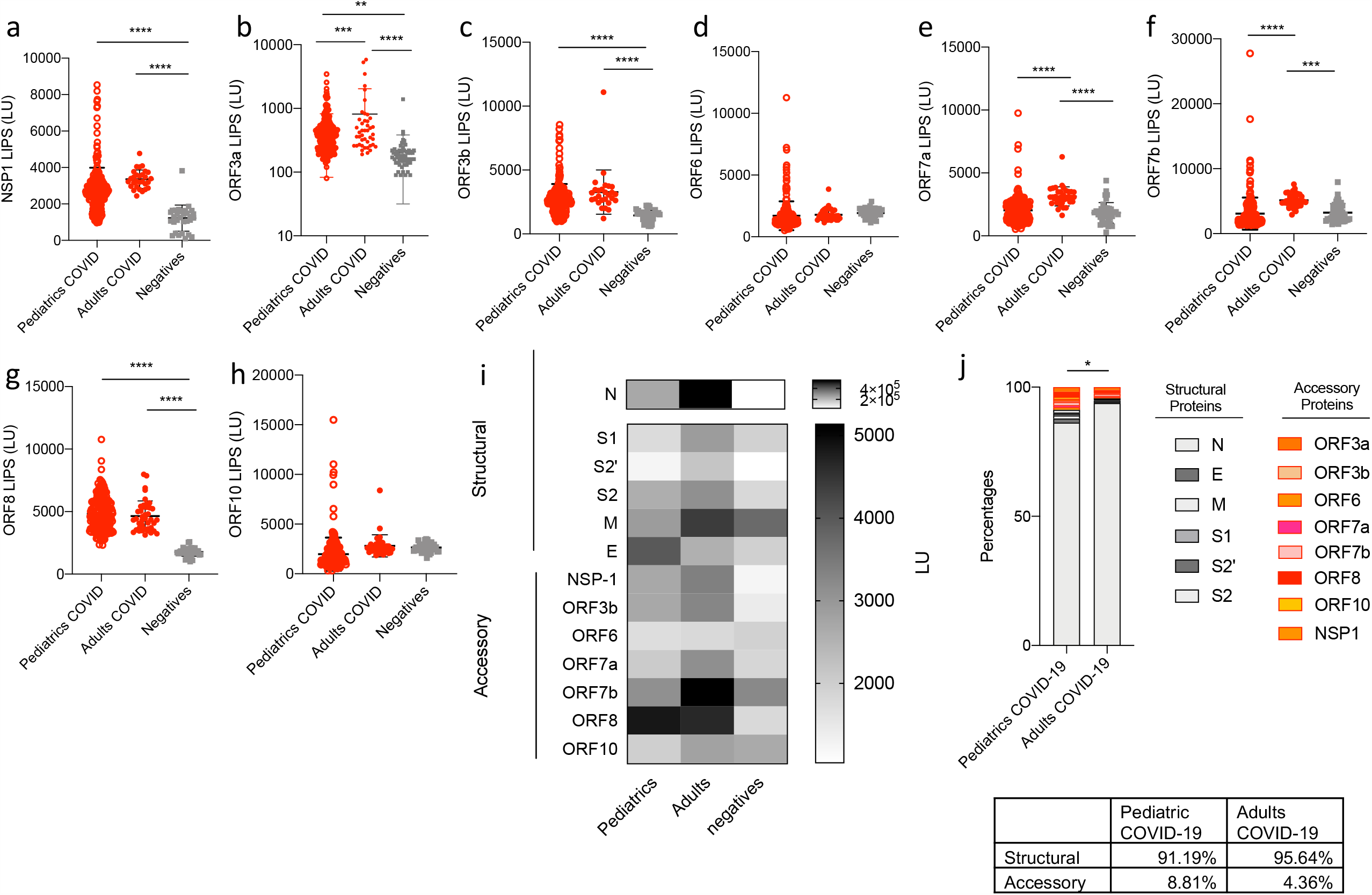
Antibody responses to SARS-CoV-2 non-structural proteins and ORFs are lower in magnitude in children than in adults with COVID-19 but represent globally a higher proportion of the SARS-CoV-2 humoral response. Antibodies against NSP1 (a) (in ORF1ab), and other ORFs (ORF3a (b), ORF3b (c), ORF6 (d), ORF7a (e), ORF7b (f), ORF8 (g) and ORF10 (h)) were measured in pediatric (n=254) and adult (n=36) COVID-19 cases and negative controls (n=33) by LIPS to cover all the ORFs of the virus. (i) A heatmap comparing the mean titres (LU) for structural (N, S, S1, S2’, S2, M, E) and accessory proteins (NSP1, ORF3a, ORF3b, ORF6, ORF7a, ORF7b, ORF10) responses in the COVID-19 pediatric and adult populations and Negatives. (j) Percentages of single antibody levels to SARS-CoV-2 antigens of the cumulative SARS-COV-2 antibody response in COVID-19 children and adults for the 14 antigens. Experiments were repeated twice. Two-sided P values were calculated using the Mann-Whitney U test. * shows statistical significance between COVID-19 patients versus negative controls. *p<0.05, **p<0.01, ***p<0.005, **** p<0,0001. Data in (a-h) represents the individual responses and mean +/- stdev, data in (i) represents mean values (LU), data in (j) represents percentages.

Cumulative SARS-CoV-2 antibody responses from COVID-19 children and adults populations were then compared as percentages of the total SARS-CoV-2 structural and accessory antibody response. The anti-N antibodies substantially dominate the SARS-CoV-2 humoral response detected by LIPS in both populations (Figure 2i-j), which is consistent with our previous findings in the adult population^14^. Due to the immunodominant effect of anti-N antibodies, we also performed analysis with or without N, both of which were highly significant (p<0.0001 Supplemental Figure 1b-e). Furthermore, representation of cumulative percentages of single specific antibodies to the global SARS-CoV-2 antibody response shows that the amount of the response towards the accessory proteins (NSP1 and ORFs) is increased over the response towards the structural ones in the pediatric COVID-19 population versus the adult COVID-19 population (8.81% versus 4.36% for the response to accessory proteins, p=0.019, Figure 2j) despite no significant differences in total IgG levels in both populations (Supplemental Figure 1a).

### Deciphering the SARS-CoV-2 antibody landscape differences in children and adults using clusters of points and principal component analysis

A cluster of points depicts each individual sample in a more complete way than a single statistical comparison, as it considers a combination of three (or more) different parameters taken together and the relevant relations of these parameters. To decipher the SARS-CoV-2 antibody landscape in children, we used relevant antibody combinations to represent the COVID-19 pediatric samples in clusters of points along with the negative and COVID-19 adult populations (Figure 3a-c and Supplemental figure 2).

**Figure 3.**
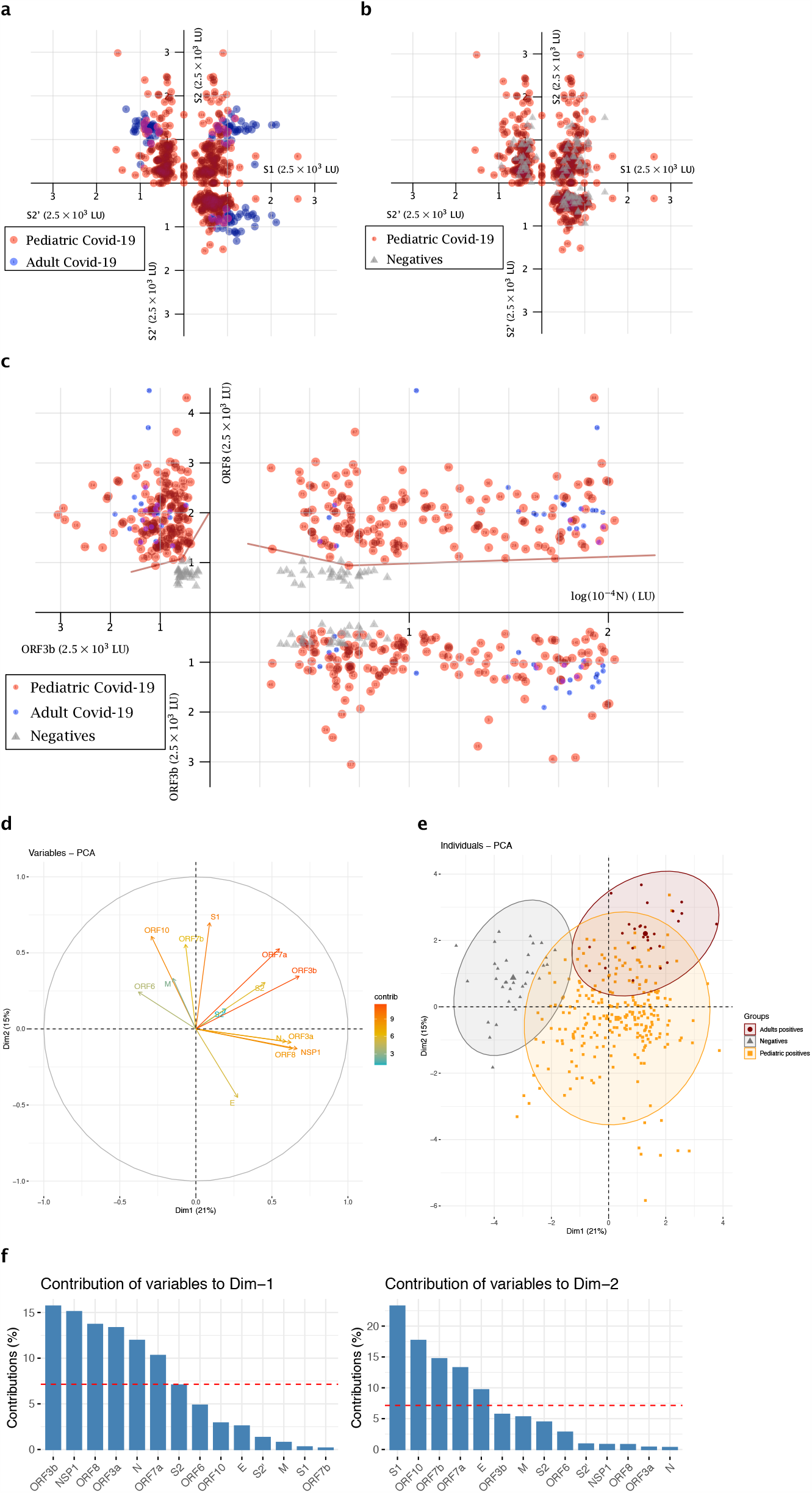
Representation of the pediatric COVID-19 population as a cluster of points for relevant antibody combinations and Principal Component Analysis (PCA). (a-b). Cluster representation of S1, S2’, S2 antibodies combination. (a) shows the pediatric COVID-19 population versus the adult COVID-19 population, (b) shows the pediatric COVID-19 population versus the negative population. (c) Cluster representation of N, ORF3b, ORF8 antibodies combination, for the pediatric COVID-19 population versus the adult COVID-19 population and the negative population. Patients are presented according to their values of SARS-CoV-2 individual LIPS antibodies as (*x, y, z*) in the space. Pediatric COVID-19 patients (n=144) are represented as red dots. COVID-19 adult patients (n=36 in (a-b) and n=24 in (c)) are represented in blue. The negative population (n=28) is represented in gray. (d-f) PCA of 14 antibodies analyzed in COVID-19 pediatric patients. Dim1 explains 21% of the variation, while Dim2 explains 15% of the variation. (d) Correlation circle and contributions. The scale of contributions is indicated (right). (e) Contribution of variables on dimensions 1 and 2. The red dashed line on the graph above indicates the expected average contribution. (f) Factorial plot of PCA on dimension 1 and 2. The plot is colored by sample types, the largest point in shape in each group is the group mean point (circle is for Adult positives, triangle for Negatives and squares for Pediatric positives).

First, the cluster representing the three antibodies to the S subunit antigens S1, S2’, S2 confirmed that the pediatric population has a S antibody profile that is more closely comparable to negative controls (Figure 3a) than an adult COVID-19 response by LIPS (Figure 3b). Further cluster analysis of antibodies to S1, S2’ with N, or other structural proteins N, M, and E reveals that the COVID-19 children population appears to be quite heterogeneous (Supplemental Figure 2a-b). Despite having a different profile than both the adult COVID-19 and the negative populations, the pediatric population cannot be clearly discriminated.

We then selected accessory protein antibodies as combinations to investigate the relevance of under-utilized markers. ORF3b and ORF8 antibodies were selected, along with N antibodies, as both were previously shown to discriminate accurately COVID-19 adults from negative controls^14^. The (N, ORF3b, ORF8) cluster of points can accurately allow the positive discrimination of the pediatric COVID-19 cases from the negatives (Figure 3c). Indeed, in the (N, ORF8; x, y) plane, the negative population is separated from the adult and pediatric positive ones by two-segments of straight lines (equations of 830*log (N) +0.3843*ORF8=4801 and -350*log (N) +1.036*ORF8=790, with all positive samples represented above or on these lines, and only one negative sample being above these lines). Then, using the (ORF3b, ORF8; y, z) plane, again, two-segments of straight lines (equations of 0.035*ORF3n+0.1334*ORF8=409.284 and 0.074*ORF3b+0.0437*ORF8=221.812) separate the negative samples from the adult and pediatric positive ones. Therefore, the (N, ORF3b, ORF8) cluster reveals that the pediatric COVID population resembles a COVID-19 adult population and can be discriminated from negative pre-pandemic controls. Importantly, this is the only combination that allowed us this discrimination, as other parameter combinations (e.g. (N, S1, S2’), (N, E, M) in Supplemental Figure 2) and combinations of antibodies to accessory proteins) were also tested and represented as clusters of points but did not discriminate pediatric samples. These data cluster analysis show that the antibody landscape of the COVID-19 children population is distinct from the adult one.

To test the hypothesis that the antibody landscape to structural and accessory viral proteins drives the distinct profile of the pediatric population, we undertook a principal-component analysis (PCA) of the 14 SARS-CoV-2 antibodies for the full data set (from Figure 1 and 2). Dimension (principal component) 1 and 2 explained respectively 21% and 15% of the total variances from all the 14 antibody types (Figure 3d-e). Accessory proteins ORF3b, NSP1, ORF8, ORF3a, ORF7a, ORF6 and ORF10 had high correlation values (Supplemental table 1), reflecting that antibodies to structural proteins do not solely drive the principal component 1. Particularly, contributions of ORF3b, NSP1, ORF8, ORF3a were the highest in Dimension 1 (Dim1, Figure 3de). Moreover, PCA showed that ORF3b and ORF7a antibodies highly contributed to the differences seen in both dimensions (Figure 3d) highlighting their importance in the serological response.

Strikingly, the PCA revealed that pediatirc COVID-19 antibody response was also intermediate between COVID-19 adults and negatives (Figure 3f). Indeed, the normal-probability representation of the 3 populations showed that only 18.9% of the pediatric patients overlapped with the ellipse of the COVID-19 adults and only 3.54% overlapped with the ellipse of the negatives (Figure 3f). Further analysis on gender, time-point, symptoms and neutralization data (PRNT90) values reported that these were not significant factors in discriminating the data (Supplemental Figure 3). Therefore, the differences in the observed SARS-CoV-2 antibody responses is primarily explained by the age and type of patients: pediatric COVID-19, adult COVID-19 or pre-pandemic negative controls.

### No difference in antibody responses between symptomatic and asymptomatic COVID-19

To assess the potential effect of antibodies to structural and non-structural proteins of SARS-CoV-2, we further stratified data (from Figure 1 and 2) into symptomatic (mild/severe) and asymptomatic for both the adult and pediatric cohorts. We found no differences in antibody responses between asymptomatic and mild COVID-19 children for all 14 antigens. In adults, we observed the same trend excluding ORF3a antibody levels which are higher for symptomatic patients (p=0.0403, Figure 4b). More importantly N, M, ORF3a and ORF7b antibody levels in asymptomatic children versus asymptomatic adults were not significantly different (p=0.5673, p=0.2669, p=0.9185 and p=0.0859 respectively, Figure 4), whilst symptomatic adults had an upregulated antibody response to these antigens compared to symptomatic children (p<0.0001 for all 4 antigens, Figure 4 and Supplemental Figure 4a).

**Figure 4.**
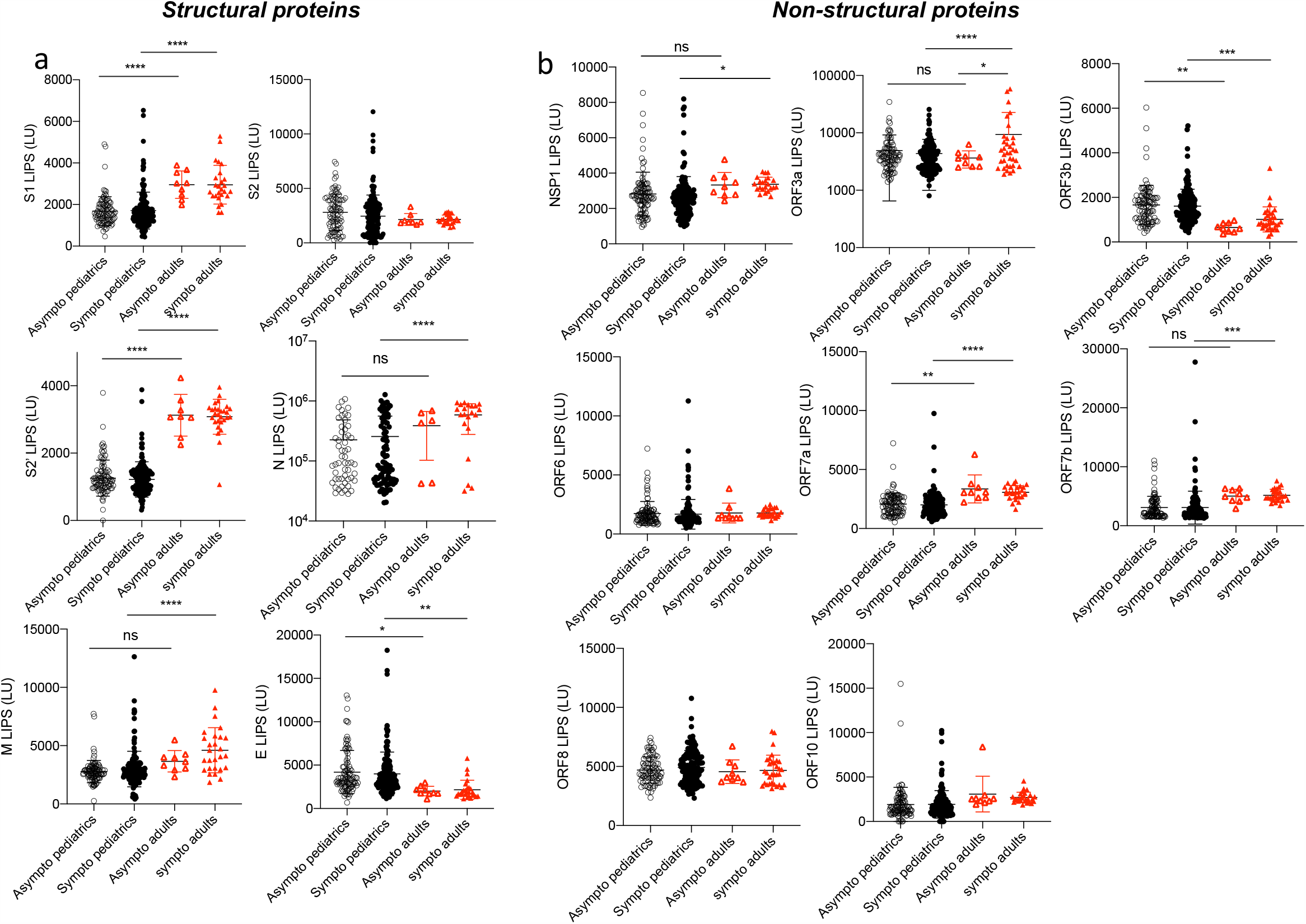
Asymptomatic and mildly symptomatic children do not display different antibody landscapes. Pediatric and adult samples were stratified according to the symptom score of the patients (asymptomatic « asympto » (pediatric COVID-19 n=98, adults COVID-19 n=9) versus symptomatic « sympto » (pediatric COVID-19 n=156, adults COVID n=27)), data from Figure 1 and 2 were analyzed according to “asympto” and “sympto”. (a) Antibodies against the SARS-CoV-2 structural proteins S1, S2, S2’, N, E, and M by LIPS. (b) Antibodies against SARS-COV-2 NSP1 (in ORF1ab), and all other ORFs (ORF3a, ORF3b, ORF6, ORF7a, ORF7b, ORF8 and ORF10). Two-sided P values were calculated using the Mann-Whitney U test. * shows statistical significance between COVID-19 patients versus negative controls. *p<0.05, **p<0.01, ***p<0.005, **** p<0,0001. All data represent individual responses and the mean +/- stdev.

### Antibody landscapes at early infection and long-term stability

We previously observed that the SARS-CoV-2 antibody responses can vary in magnitude and specificity in adults between acute and convalescent to memory time-points^14^. To study the effect of time in the pediatric COVID-19 population, we stratified pediatric responses of all 254 samples (Figures 1 and 2) by early (<d14) versus later (≥d14) time-points (Figure 5). S2, N and ORF7a specific antibodies were significantly increased after day 14 post symptom onset. In contrast, ORF3b and ORF7b antibodies elicited a higher antibody response prior to day 14 (Figure 5b). Finally, responses to structural proteins S1, S2, M and E and accessory proteins NSP1, ORF3a, ORF6 and ORF8 were comparable before and after day 14 (Figure 5ab).

**Figure 5.**
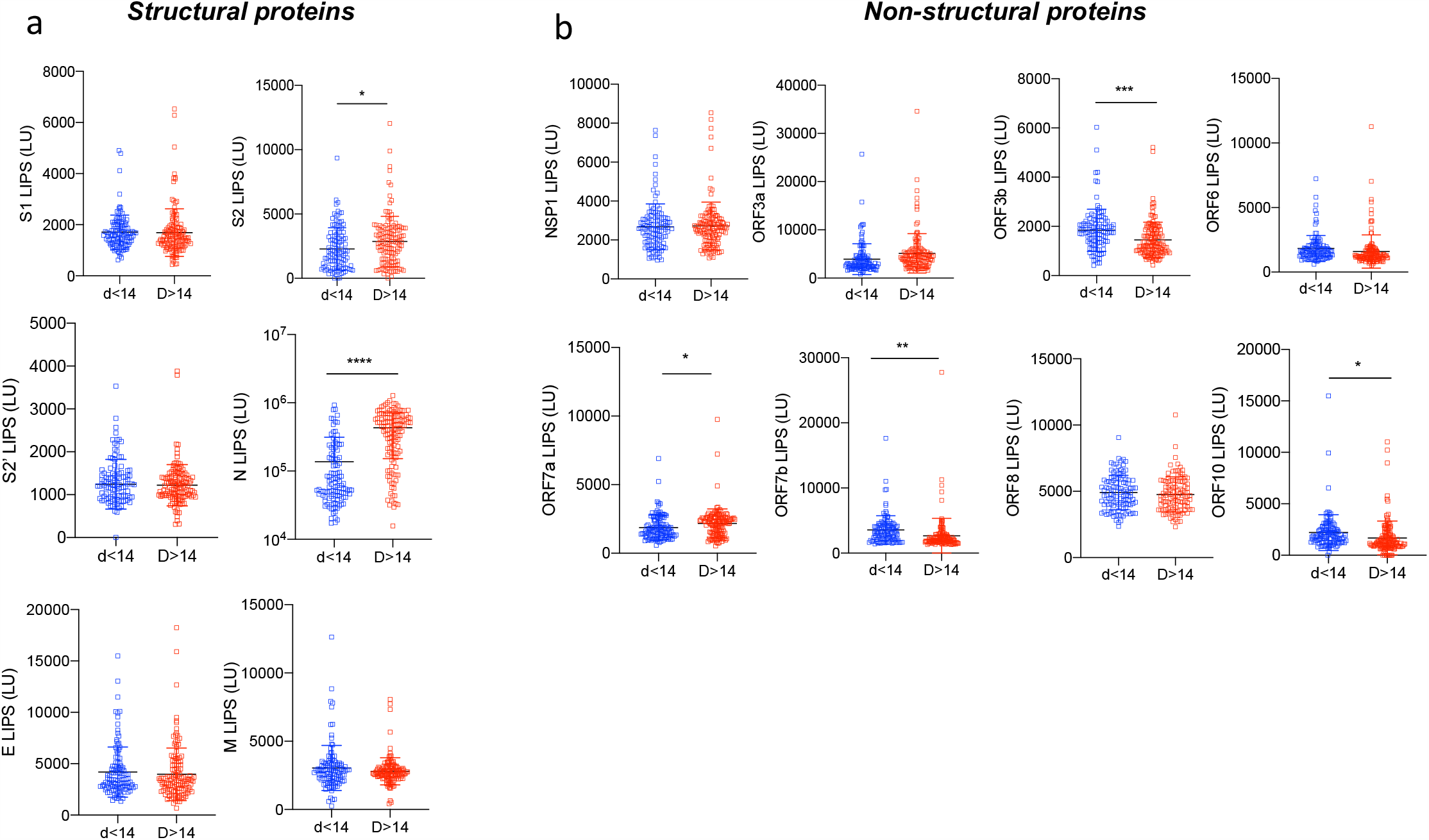
A unique antibody landscape is specific of early time-point samples (< day 14). Pediatric samples were stratified according to the time-point of collection, and data from Figure 1 and 2 were analyzed according to acute (<day 14, n=119) and later time-points (≥day 14, n=135). (a) Antibodies against the SARS-CoV-2 structural proteins S1, S2, S2’, N, E, and M by LIPS. (b) Antibodies against NSP1 (in ORF1ab), and all other ORFs (ORF3a, ORF3b, ORF6, ORF7a, ORF7b, ORF8 and ORF10). P values were calculated using the student t test. * shows statistical significance between acute time-point pediatric COVID-19 patients versus late time-point pediatric COVID-19 patients. *p<0.05, **p<0.01, ***p<0.005, **** p<0,0001. All data represent individual responses and the mean +/- stdev.

To further confirm the stability of SARS-CoV-2 specific antibodies we used 146 longitudinal paired samples of 58 pediatric patients that had either 2, 3 or 4 blood draws (Figure 6a). The time-frame of sampling ranged from 0 to 206 days post-symptom onset, with the majority of samples from <14 days (n=63), or long term memory samples after day 60 (n=58) (Figure 6b). Using a linear mixed effects model, we determined that antibody responses to structural proteins S1, S2, S2’, M and E were stable over time, whereas N was significantly increased (p<0.001) (Figure 6c). Furthermore, antibodies towards non-structural proteins NSP1, ORF3a, ORF3b and ORF7a also significantly increased over time (p<0.001, p=0.001, p=0.027 and p=0.002 respectively), whilst ORF6, ORF8 and ORF10 were stable (Figure 6c). Only ORF7b antibody response significantly decayed longitudinally at a slow rate (p<0.001, Figure 6c). In order to determine whether the slope of each serological marker could inform the disease outcome we compared asymptomatic and symptomatic patients but no significant differences were found (p>0.05 for all) (Supplemental figure 4).

**Figure 6.**
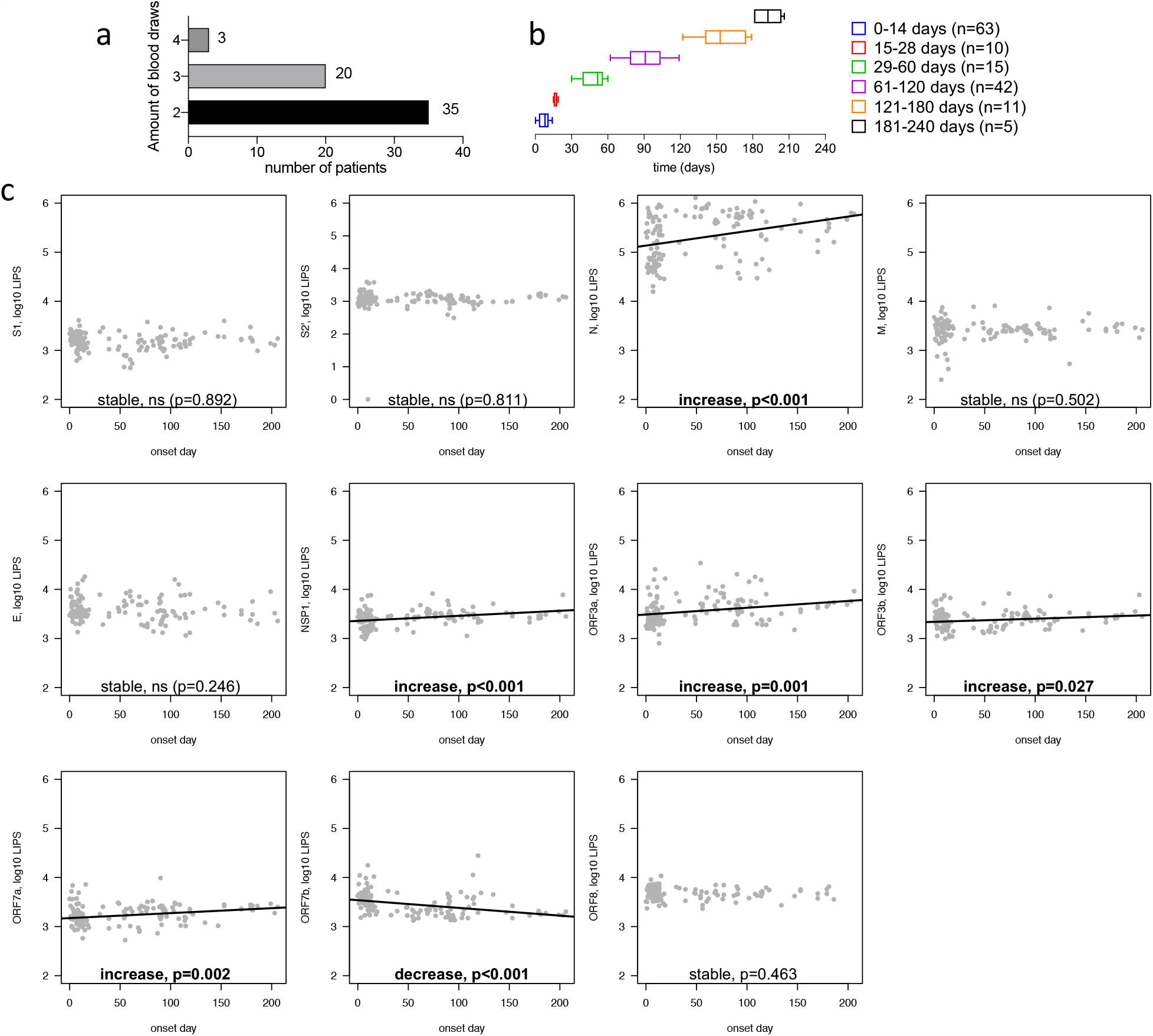
Longitudinal stability of antibody responses for structural and non-structural SARS-CoV-2 proteins in COVID-19 children. (a) Number of longitudinal patients with either 2, 3 or 4 blood draws from 58 pediatric COVID-19 cases. (b) Sample collection time-line (days post infection). (c) A linear trend on log_10_ LIPS values was fitted for longitudinal samples for S1, S2’, N, M E, NSP1, ORF3a, ORF3b, ORF7a, ORF7b, ORF8 (n=58 pediatric COVID-19 patients).

### A distinct antibody landscape may impact IFNα levels

Severe COVID-19 disease is associated with low IFNα responses in adults which has been linked to the type-I IFN down-regulation roles of ORF3b, ORF7a and ORF6^15-17^. Antibodies to these 3 proteins in a cluster of points (ORF3b, ORF7a, ORF6 as *x,y,z*), show that children have a heterogenous humoral profile towards these 3 type-I IFN down-regulators (Figure 7a). To assess the IFNα response a quantitative ELISA was conducted on plasmas collected before day 7 in children (n=48) and adults (n=18) (Figure 7b). We observed a significant decrease of acute IFNα levels in children compared to the adult samples (p=0.0165, Figure 7b), with only 3 pediatric samples showing a detectable IFNα level. To further investigate the antibody profiles with IFNα responses, the total antibody response of these 3 IFNα producing children compared to the non-responder children (n=45), we plotted the average antibody responses to all 14 antigens for each group (Pediatric IFNα^-^ versus Pediatric IFNα^+^, average: Figure 7c, and individuals: Supplemental figure 6). We found a significant difference in the antibody distribution in the 3 IFNα producing children versus the IFNα non-producing children (IFNα^+^ versus IFNα^-^, p<0.0001), with IFNα^+^ children having increased ORF6, ORF7a and ORF3b antibodies, and lower ORF8 and E antibody levels. Notably ORF6 antibody levels were particularly higher in one IFNα^+^ asymptomatic child with low viral loads (Figure 7d).

**Figure 7.**
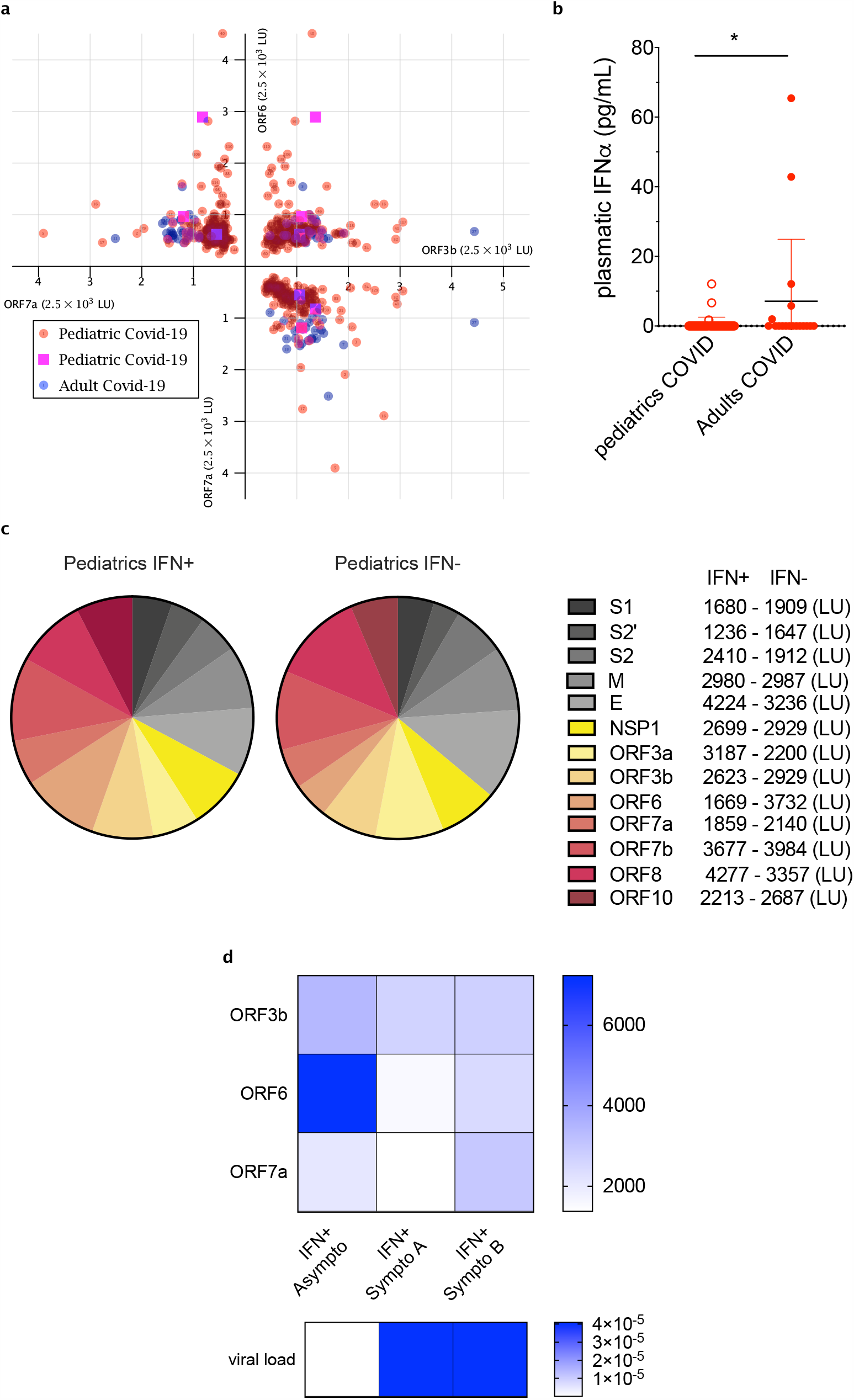
IFN-α producing pediatric patients display a different landscape of antibody response to SARS-CoV-2 accessory proteins. (a) Cluster representation of antibodies to the accessory proteins ORF3b, ORF7a, ORF6 (x, y, z) for the COVID-19 pediatric samples (red, n= 144) versus the COVID-19 adult samples (blue, n= 27). (b) Plasma IFN-α concentrations (pg/ml) in pediatric (n=48) and adult COVID-19 cases (n=18) early timepoint samples (< day 7). Data represents individual responses and the mean +/- stdev. (c) Pie charts of the cumulative antibody responses to the relevant SARS-CoV-2 structural and non-structural protein antigens (excluding N) in COVID-19 pediatric cohort stratified (positive/negative) by their IFN-α responses. (d) Individual data for IFN-α^+^ pediatric cases viral loads by RT-PCR, ORF3, ORF6, and ORF7b LIPS LU. P values were calculated using Chi-squared test between the mean of IFNα-pediatric COVID-19 patients (N=45) and the IFNα^+^ (N=3). ns, p=0.0591 *p<0.05, **p<0.01, ***p<0.001, **** p<0.0001.

## Discussion

Young children account for only a small percentage of reported and medically attended COVID-19 infections^5^, which is unlikely to be completely explained by reduced exposures and school closures. This difference is likely contributed to by differences in host responses between children and adults. We present herein the most comprehensive study to date of the magnitude, specificity and duration of SARS-CoV-2 specific antibodies in children.

While the understanding of immunity to COVID-19 is growing at a fast pace, information on the pediatric population remains limited largely due to their asymptomatic and mild illness compared to the adult population. Furthering our understanding of the immune mechanisms that lead to this mild clinical presentation could represent new alternative therapeutics, prevention methods or improved diagnostics. Our work enables unique serological insights on the long-term response and asymptomatic infections as Hong Kong’s pandemic control strategy has intensive testing, contact tracing and isolation of all COVID-19 cases including asymptomatic and mild cases with longitudinal follow up. This has provided us the opportunity to investigate a cohort of children with both symptomatic and asymptomatic children. We describe the antibody diversity between day 0 and day 206 post-symptom onset, and reveal major differences in the antigens targeted by the humoral immune response of COVID-19 children compared to adults which may indicate differences in viral protein propagation kinetics, pathogenesis and IFN immune evasion.

Importantly, we report that the proportion of the antibody response targeting the accessory proteins is significantly increased in children versus adults. Antibodies to the N structural protein largely dominate the humoral immune response both in children and adults, though the total magnitude of the N-specific antibodies in children is substantially lower than adults. Visualization of N antibody levels in our cluster representations clearly show that only half of the pediatric cases have comparable levels of these antibodies to adults. Others have also reported a reduced magnitude of N antibodies in children and they suggested that this observation could be related to a lower release of N proteins related to lower replication in children^18^. On the contrary, our data show that children produce antibodies to some accessory proteins (namely NSP1, ORF3b and ORF8) at similar levels to adults, and to structural protein E in higher proportions than adults, the latter notorious for high turnover due to its pivotal role in viral propagation (reviewed in^19^). Therefore, these accessory proteins are not being released to a lesser extent in children but may reflect different virus pathogenesis in children compared to adults. Viral loads have been shown to be comparable in children and adults, which may reflect similar levels of viral replication^2^.

For the Spike subunits antibodies, the (S1, S2’, S2) cluster reveals that the children population resembles a negative pre-pandemic population and not a COVID-19 adult one. A recent study describes a lower anti-S IgG, IgM, IgA in the pediatric population which correlates with our findings ^20^. One explanation on the clinical difference between children and adults raise that the pre-existing immunity against seasonal human coronaviruses (HCoVs) that cross-reacts with SARS-CoV-2 is higher in children, as they have a higher infection rate of seasonal HCoVs than adults^21^. Individuals exposed and unexposed to SARS-CoV-2 have cross-reactive antibodies against the proteins of SARS-CoV-2 and seasonal HCoVs^22,23^. Moreover because circulating HCoVs have a higher homology to SARS-CoV-2 structural proteins than non-structural proteins (if they exist)^24,25^, we would expect a higher cross-reactivity for structural proteins based on pre-existing immunity. SARS-CoV-2 infection back-boosts antibodies against conserved epitopes, including the relatively conserved fusion peptide of the Spike S2 subunit^22,23^. In our hands, COVID-19 children and adults had comparable levels of S2 antibodies, contrary to S1 and S2’, which shows a possible effect of pre-existing HCoVs immunity for more conserved domains of S such as S2.

Our observations of lower Nucleocapsid and Spike antibodies in COVID-19 children may indicate that there may be lower sensitivity of serological detection for SARS-CoV-2 when using assays based on S and/or N alone, leading to an underestimation of SARS-CoV-2 exposed children. S antibodies have been reported in lower magnitude in the majority of mild adult infections, with higher levels being produced in severe cases^26^, which is consistent with our data on low S antibody levels in children which were also asymptomatic or mild clinical scores. Low antibody levels and low affinity have been associated with Antibody Dependent Enhancement by facilitation of viral uptake by host cells^27^, however yet no definitive evidence of ADE for SARS-CoV-2 neither in adults nor in children has been brought forward^28^, but warrants further investigation.

The plane (ORF3b/ORF8) in the cluster of points (N, ORF3b, ORF8) reveals that children samples have specific combinatory values of these two antibodies that is consistent with adult populations, and that makes them distinguishable from the negatives. Similar to NSP1, we report the proportion of all non-structural antibodies makes a greater contribution of the SARS-CoV-2 response in children than in adults. The Principal Component Analysis of our dataset confirmed further the importance of antibodies to accessory proteins in characterising the pediatric samples. Whether these antibodies to accessory proteins play a role in the virus infectivity or in the pathogenesis of the disease and in the milder outcome of SARS-CoV-2 infection in children presents further questions for investigations.

ORF3b, ORF7a and ORF6 proteins have been previously reported to play a role in cellular type-1 IFN down-regulation^15-17^. The cluster representation of ORF3b, ORF7a and ORF6 antibodies shows a different pattern between adult and children population. Furthermore, the PCA revealed that ORF7a and ORF3b contributed highly to component 1 and 2 (Dim1 and Dim2) which accounted for 21% and 15% of the variances observed respectively, pointing to a potential pivotal role of these antigens. In all COVID-19 infected children or adults tested at early timepoints of infection (< day 7), the majority did not elicit a detectable IFNα response, in line with previous findings^30^, but overall children IFNα responses were significantly lower than the adult ones. Amongst the IFNα responders and non-responders in children a different antibody landscape suggests possible functions of these markers in counteracting viral IFN down-regulation, with ORF6 antibody responses doubled in IFNα^+^ children. Bastard *et al*., reported anti-type I interferon auto-antibodies in a subset of severe COVID-19 patients^9^. Moreover, a recent study has linked high levels of auto-immunity with COVID-19 severe cases in adults^31^. As children are less predisposed to auto-immunity than adults^32^, it is possible this contributes to their milder clinical presentation along with the diversified antibody landscape observed in our study.

We report in children diverse antibody profiles in early versus late samples and the maintenance or increase of all antibodies to structural and accessory proteins, except ORF7b antibodies, for at least 6 months post-infection. Many factors play a role in antibody long-term persistence, such as antigen release, antigen presentation, induction of a germinal centre reaction and a memory B cell pool^33^. Additional studies on viral proteins release, their roles and their specific B cells are needed to fully understand the antibody landscape in children.

Our cohort did not include any case of Multi-inflammatory System in Children (MIS-C). Although one study reported that no distinct antibody response was observed between MIS-C and mild or asymptomatic children^18^, it only measured S and N antibodies. Therefore it would be of interest to study the whole spectrum of antibodies in this population and particularly those targeting accessory proteins. In our hands, symptomatic children have significant differences in antibody levels versus symptomatic adults only for selected antibodies (N, M, ORF3a and ORF7b) which suggests that these markers could play a role in infection control or infectivity.

It is possible that the interest for antibodies to SARS-CoV-2 internal proteins will grow with the rollout of sub-unit Spike only vaccines, in order to allow the distinction between SARS-CoV-2 past exposure and vaccination in specific populations and to create an estimated date of exposure given the unique rate of waning of different specificities. The emergence of certain viral mutants such as the ORF8 truncations^34^ or recent ORF3b deletions^35^ could modify the contributions of certain ORFs, their antibodies responses could also be used for epidemiology studies on the insurgence of different strains of the virus.

In conclusion, we report the description of a more diversified antibody landscape in the COVID-19 children population compared to adults, with an increased and sustained humoral response to all accessory proteins of the virus. This study of antibody spectrum provides insights into the importance of the breadth of responses and how it differs between children and adult that have diverse outcomes of infection, and could inform improved SARS-CoV-2 diagnostics for the pediatric population.

## Methods

### Patients and samples collection

Our study enrolled a total of 122 children patients and 36 adult patients based on recruitment of available patients with RT-PCR confirmed COVID-19 infection in Hong Kong. We used a total of 254 COVID-19 children plasma samples including 146 longitudinal samples from 58 subjects with 2 to 4 sampling time points, and 119 early time-points samples (< day 14). Samples were used from children (mean±stdev: 39±47 days, range: 0-206 days) and adults (mean±stdev: 54±20 days, range: 24-123 days), with the sample day was defined as day post-symptom onset or RT-PCR confirmation for asymptomatic cases through contact tracing or quarantine. For measurement of IFNα, an extra set of 18 COVID-19 adult patients sampled prior to day 7 was used in comparison to 48 samples that were collected prior to day 7 in the COVID-19 pediatric cohort. The COVID-19 patient study was approved by the institutional review board of the respective hospitals, viz. Kowloon West Cluster (KW/EX-20-039 (144-27)), Kowloon Central / Kowloon East cluster (KC/KE-20-0154/ER2) and HKU/HA Hong Kong West Cluster (UW 20-273, UW20-169), Joint Chinese University of Hong Kong-New Territories East Cluster Clinical Research Ethics Committee (CREC 2020.229). All of patients provided informed consent.

The negative control plasma samples used in this study were from Hong Kong blood donors collected from June to August 2017 (prior to the emergence of COVID-19), used a total of 33 plasma samples including negative pediatric samples (n=20) and negative adult samples (n=13). The collection of negative control blood donors was approved by the Institutional Review Board of The Hong Kong University and the Hong Kong Island West Cluster of Hospitals (approval number: UW16-254). Plasma samples were collected from heparinized blood. All samples from COVID-19 patients or negative controls were heat-inactivated prior to experimental use at 56°C for 30 minutes. Details on the sample cohort are presented in Table 1.

### SARS-CoV-2 cloning and (Ruc)-antigen expression

Based on previous studies describing the structure of the SARS-CoV-2 genome^25,36^, an extensive panel of 14 proteins (S1, S2, S2’, E, M, N, NSP1, ORF3a, 3b, 6, 7a, 7b, 8, 10) was chosen for antibody testing by LIPS. Primers and cloning for the amplification of SARS-CoV-2 proteins were as previously described^14^. Constructs with pREN2-Renilla luciferase plasmid containing the SARS-CoV-2 antigen of interest were transfected into Cos1 cells and prepared as previously described^14^.

### Measurement of antibody responses using LIPS

The LIPS assays were performed following the protocol of Burbelo *et al*., with the following modifications^36^, as previously described^14^. Briefly, (Ruc)-antigen (at an equal concentration for each antigen at 10^7 per well) and plasma (heat inactivated and diluted 1:100) were incubated for 2 hours with shaking at 800rpm. Ultralink protein A/G beads (Thermo-Fisher) were added to the (Ruc)-antigen and serum mixture in a 96-deep-well polypropylene microtiter plate and incubated for 2 hours with shaking at 800rpm. The entire volume was then transferred into HTS plates (Millipore) and washed as previously described. The plate was read using QUANTI-Luc Gold substrate (Invivogen) as per manufacturer’s instructions on a Wallac MicroBeta JET luminometer 1450 LSC & Luminescence counter and its software for analysis (PerkinElmer). Experimental controls include no plasma blank wells with (Ruc)-antigens and negative control serum from healthy donors plasma collected prior to the COVID-19 pandemic. The background corresponds to the LU signal from each Ruc-fusion antigen with protein A/G and substrate with no plasma.

### Enzyme-linked immunosorbent assay

Total IgG were measured in plasma samples using the Total human IgG ELISA kit (Thermo-Fisher) at a final dilution of 1:500,000 according to manufacturer’s instructions. IFN-α was measured in plasma samples using the Human IFN-α Platinum ELISA kit (Invitrogen) at a dilution of 1:5 according to manufacturer’s instructions.

### Clusters of points

The SARS-CoV-2 antibodies dataset has been treated through the free software ConTeXt, with LuaMetaTeXengine (version 2020.05.18) developed by Hans Hagen <u>(http://www.pragma-ade.nl)</u> which uses TeX, Metapost and Lua to obtain the 3D clusters of points shown in Figure 3abc, Figure 7c and Supplemental Figure 2. For clarity, only the first 144 COVID-19 pediatric samples of the dataset are represented in the clusters of points, along with n=36 COVID-19 adult samples and n=28 negatives.

In the cluster (N, ORF3b, ORF8), the equations of the red lines are: (1) in the plane (N, ORF8) :830*log (N) +0.3843*ORF8=4801 and -350*log (N) +1.036*ORF8=790, and (2) in the plane (ORF3b, ORF8): 0.035*ORF3b+0.1334*ORF8=409.284 and 0.074*ORF3b+0.0437*ORF8=221.812. These straight lines allow the most accurate discrimination between negative controls and positive adult populations.

### Principal component analysis

The LU for 14 antigens were log-scale transformed (the negative and zero values in the data set were replaced by 1) prior to PCA analysis. The missing values in the dataset were estimated by a probabilistic model ^37^. The probabilistic model is tolerant to amounts of missing values between 10% to 15% which is fit for our data. The missing data was estimated using pcaMethods (version 1.80.0)^38^. The completed data were standardized (scaled) before input in standard PCA (using *FactoMineR* (version 2.4)^39^.. The PCA results were extracted and visualized using factoextra (version 1.0.7)^40^.

### Statistics and Reproducibility

GraphPad Prism version 8 software (San Diego, CA) was used for statistical analysis. All experiments were repeated twice independently. Antibody levels are presented as the individual responses and geometric mean +/- standard deviation (stdev). Ordinary one-way ANOVA with Tukey’s multiple comparison test were performed to compare the pediatric, adult and negative populations in Figures 1 and 2, and the early and late samples in Figure 5. For Figure 2j, percentages were calculated by dividing each mean antibody value by the sum of the total antibody responses, and compared using a Chi-square test between the “observed” (pediatric) versus “expected” (adult) distributions.

For Figure 6c and Supplementary figure 3, a linear mixed effects model was fitted to account for correlated responses for the longitudinal samples dataset. Log_10_ LIPS was used for the analysis (as dependent variable) to reduce the impact of extreme values/non-normality. For Supplementary figure 1, the distributions showed in the pie-charts were compared using a Chi-square test between the “observed” (pediatric) versus “expected” (adult) distributions.

## Data availability statement

The data that support the findings of this study are available from the corresponding author upon request.

## Data Availability

Data availability statement
The data that support the findings of this study are available from the corresponding author upon request.

## Acknowledgements

The authors thank the patients and their families for their participation, and are grateful to the hospital staff, clinicians and nurses, particularly Karen YS Yui, for sample coordination. We thank Professor JT Wu, Dr Mahen RP Perera and Dr Kathy Leung for providing donor plasma controls. This study was partly supported by the Theme based Research Grants Scheme (T11-712/19-N), Health and Medical Research Fund (HMRF COVID-190115 and COVID-190126), National Institutes of Allergy and Infectious Diseases, National Institutes of Health (USA) (contract HHSN272201400006C).

## Ethics declaration

The COVID-19 patient study was approved by the institutional review board of the respective hospitals, viz. Kowloon West Cluster (KW/EX-20-039 (144-27)), Kowloon Central / Kowloon East cluster (KC/KE-20-0154/ER2) and HKU/HA Hong Kong West Cluster (UW 20-273, UW20-169), Joint Chinese University of Hong Kong-New Territories East Cluster Clinical Research Ethics Committee (CREC 2020.229). All of patients provided informed consent. The collection of plasma from blood donors serving as controls was approved by Institutional Review Board of The Hong Kong University and the Hong Kong Island West Cluster (UW16-254).

