## Supplementary material for "The SARS-CoV-2 antibody landscape is lower in magnitude for structural proteins, diversified for accessory proteins and stable long-term in children": Supp figures

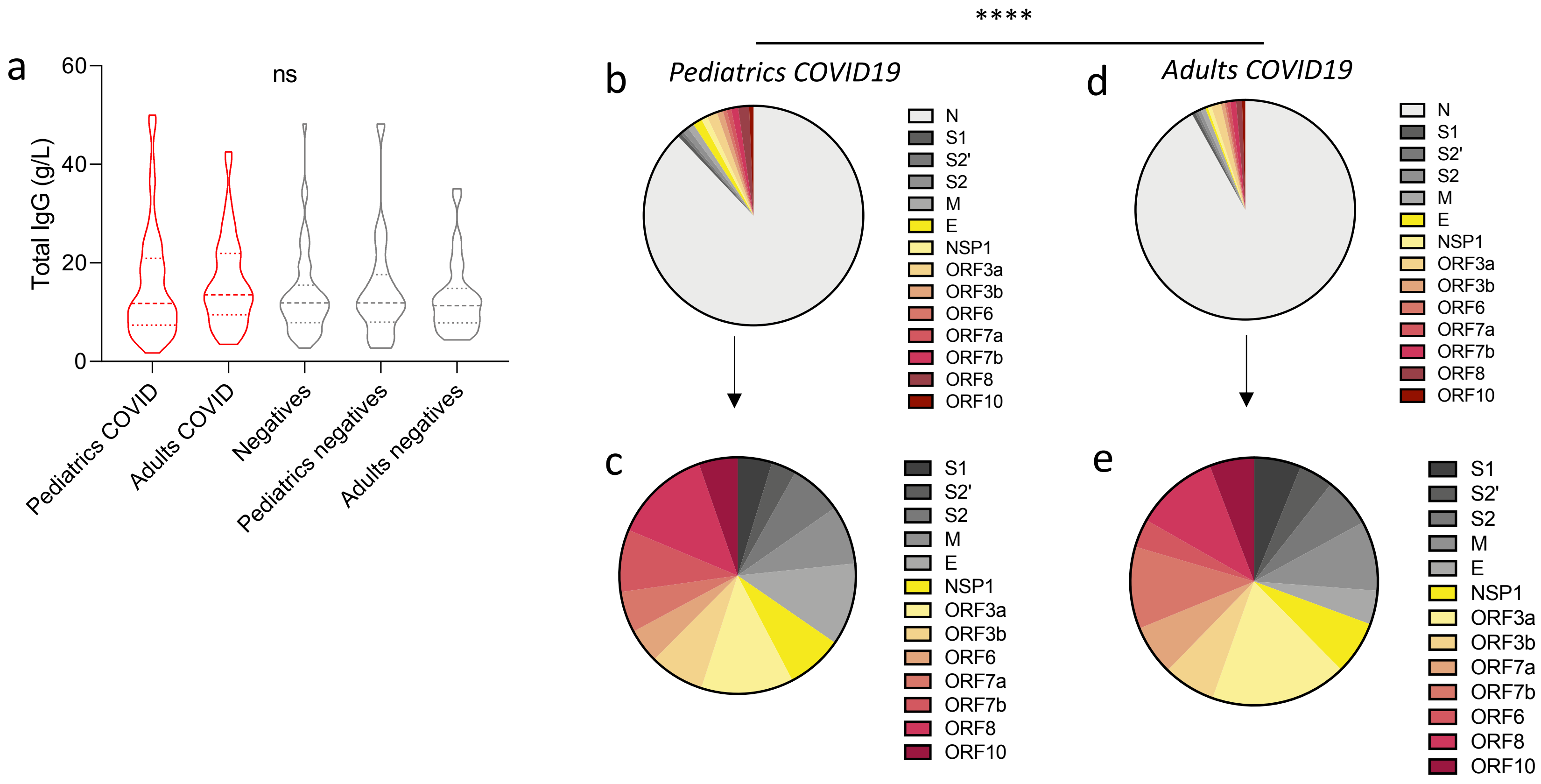


**Supplementary figure 1.** (a) Levels of total IgG (g/L) in plasma measured by ELISA in the pediatrics COVID (n=254), adults COVID (n=36), negatives (n=33), and pediatrics negatives (n=20), adults negatives (n=13). (b-e) Pie charts of the cumulative antibody responses to the 14 relevant SARS-CoV-2 structural and non-structural protein antigens in COVID-19 children (b, c) and adults (d, e). Due to the differences in scale between N and all other 10 antigens, a second pie-chart excluding N responses was done to focus on other antibody responses (c, e). * shows statistical significance. Two-sided P values were calculated using a Chi-square test between the "observed" (pediatric) versus "expected" (adult) distributions. **** p<0,0001.

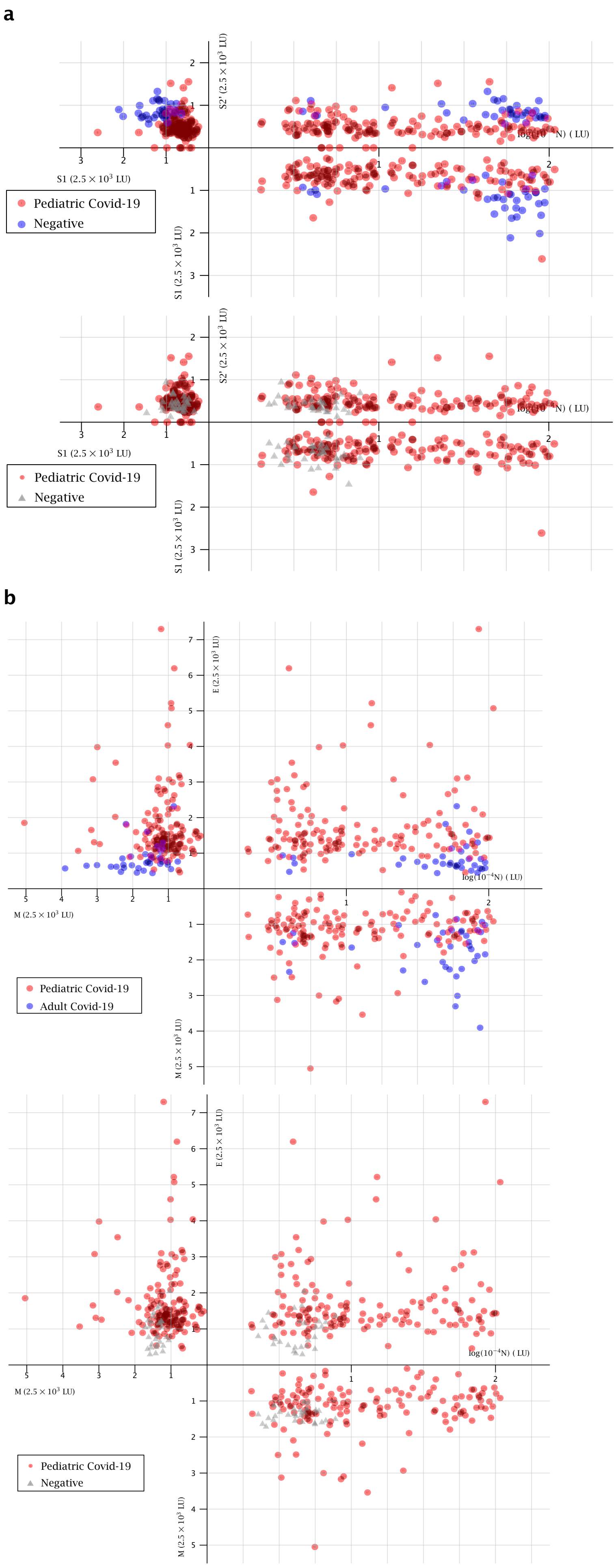
a
S2' (2.5 × 10³ LU)
log(10⁻⁴N) ( LU)
S1 (2.5 × 10³ LU)
Pediatric Covid-19
Negative
S2' (2.5 × 10³ LU)
log(10⁻⁴N) ( LU)
S1 (2.5 × 10³ LU)
Pediatric Covid-19
Negative
b
E (2.5 × 10³ LU)
log(10⁻⁴N) ( LU)
M (2.5 × 10³ LU)
Pediatric Covid-19
Adult Covid-19
E (2.5 × 10³ LU)
log(10⁻⁴N) ( LU)
M (2.5 × 10³ LU)
Pediatric Covid-19
Negative

**Supplemental Figure 2. Representation of the pediatric COVID-19 population as a cluster of points**. (a) N, S1, S2'antibodies combination for the pediatric COVID-19 population versus the adult COVID-19 population (top) and for the pediatric COVID-19 population versus the negative population (bottom). (b) N, M, E antibodies combination for the pediatric COVID-19 population versus the adult COVID-19 population (top) and for the pediatric COVID-19 population versus the negative population (bottom). Patients are presented according to their values of SARS-CoV-2 individual antibodies as ($x$, $y$, $z$) in the space. COVID-19 children patients (n=144) are represented as red dots. COVID-19 adult patients (n=36) are represented in blue. The negative population (n=28) is represented in gray.

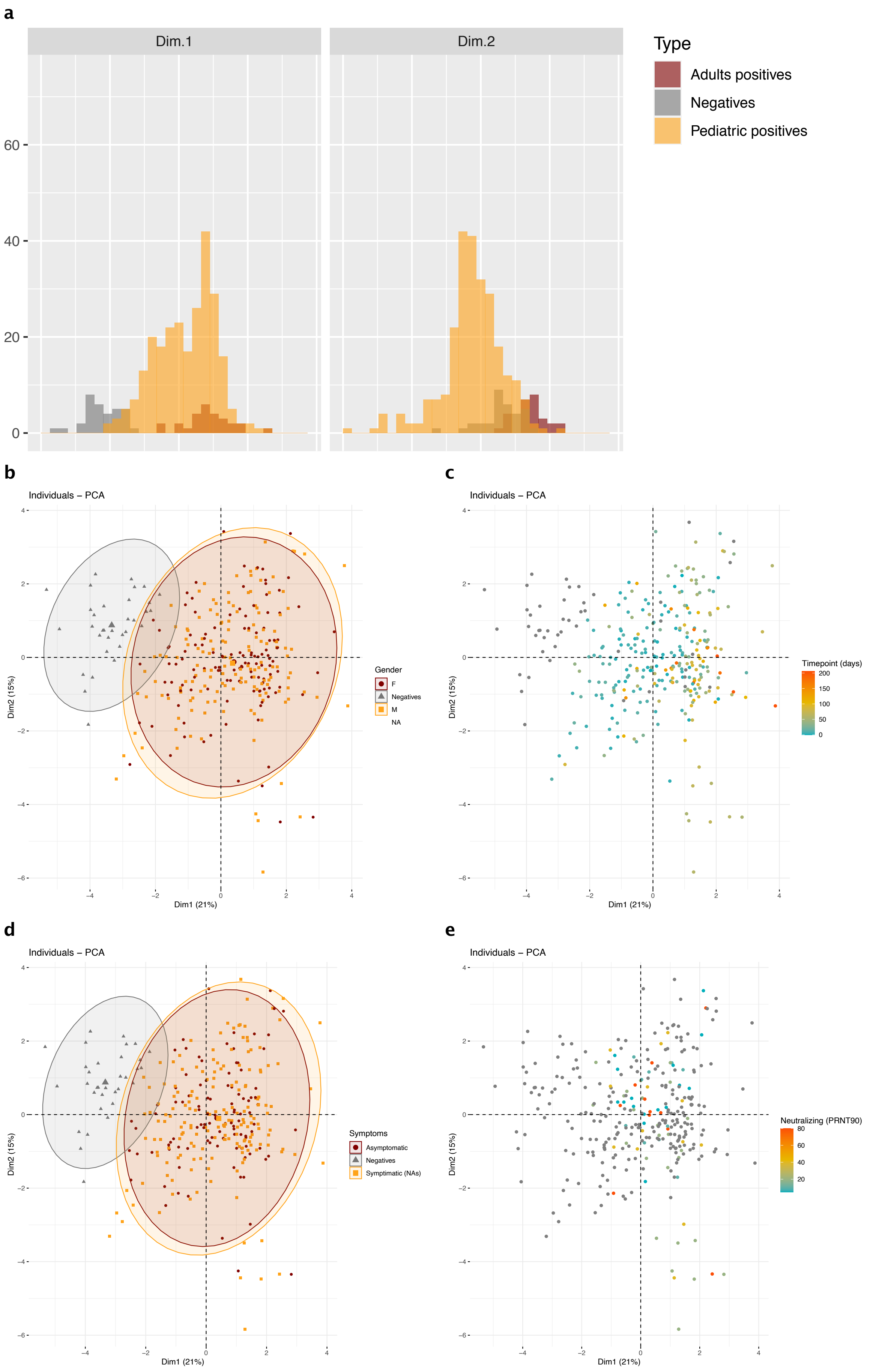
a
Dim.1
Dim.2
60
40
20
0
Type
Adults positives
Negatives
Pediatric positives
b
Individuals – PCA
Gender
F
Negatives
M
NA
Dim1 (21%)
Dim2 (15%)
c
Individuals – PCA
Timepoint (days)
200
150
100
50
0
Dim1 (21%)
Dim2 (15%)
d
Individuals – PCA
Symptoms
Asymptomatic
Negatives
Symptimatic (NAs)
Dim1 (21%)
Dim2 (15%)
e
Individuals – PCA
Neutralizing (PRNT90)
80
60
40
20
Dim1 (21%)
Dim2 (15%)

**Supplemental Figure 3. Principal Component Analysis**. (a) Distributions of the Principal Components (Dim1 and Dim2) colored by sample-type: Pediatric positives, Adult positives, Negatives. (b-e) Factorial plot of PCA on dimension 1 and 2 for gender (b), time-point (c), symptoms (d) and neutralization data (PRNT90) (e).

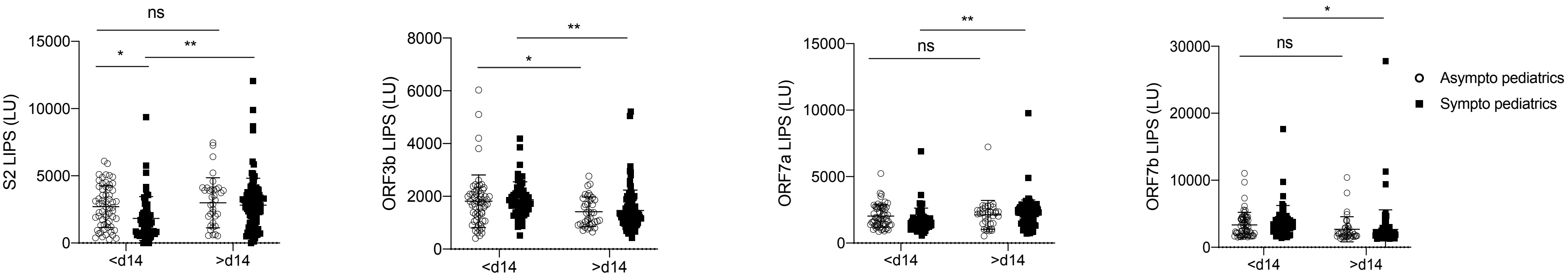


**Supplemental Figure 4.** Stratification of S2, ORF3b, ORF7a and ORF7b antibody levels measured by LIPS according to symptoms and time-point of sampling (prior day 14 or after day 14). Two-sided P values were calculated using the Mann-Whitney U test. * shows statistical significance between symptomatic and asymptomatic samples or between early and late samples. *p<0.05, **p<0.01.

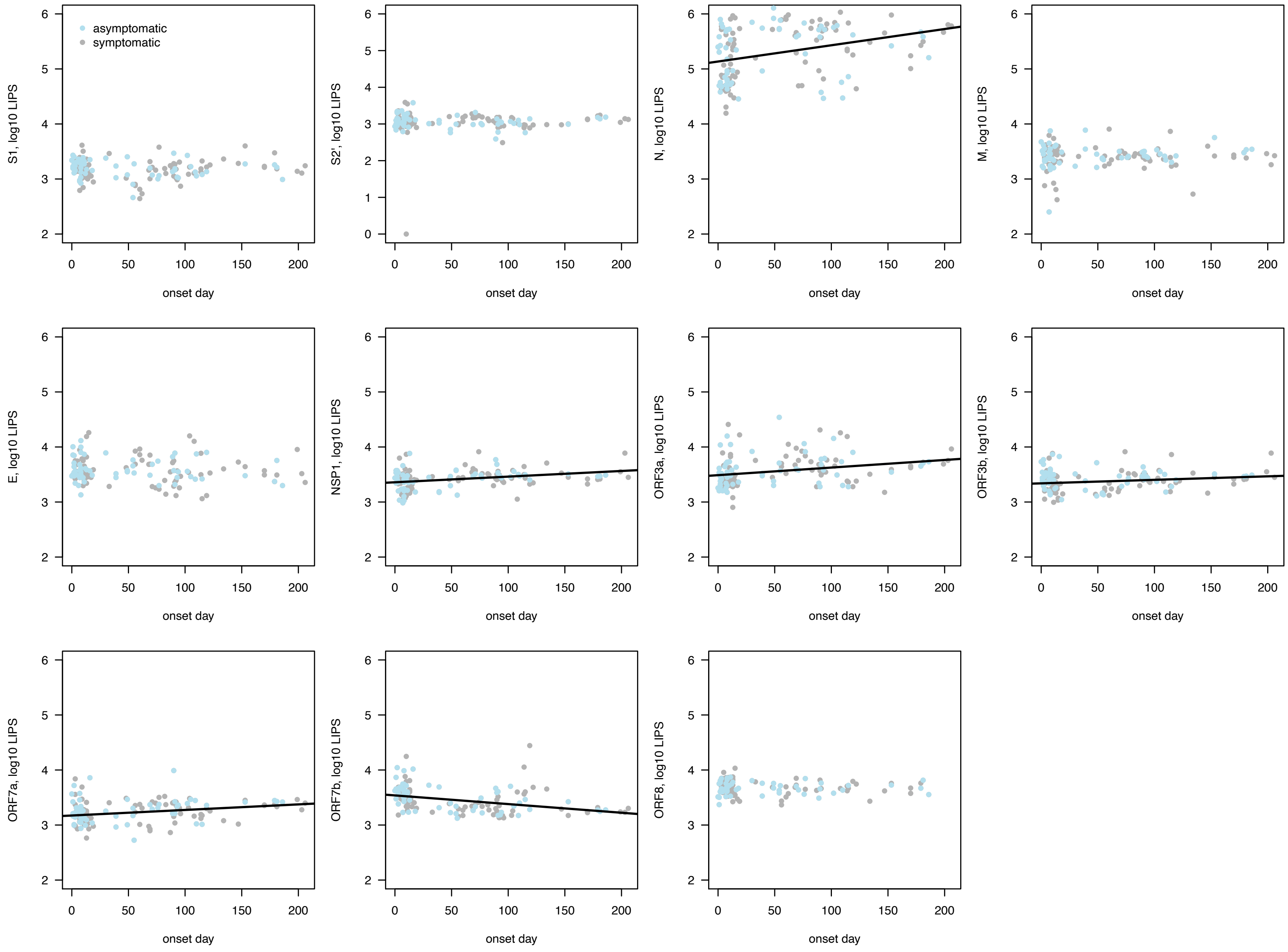


**Supplemental Figure 5. Longitudinal antibody responses in asymptomatic and symptomatic COVID-19 children.** Longitudinal analysis of 11 relevant antibody responses for structural (S1, S2', N, M, E) and non-structural (NSP-1, ORF3a, ORF3b, ORF7a, ORF7b, ORF8) SARS-CoV-2 proteins in asymptomatic (light blue) and symptomatic (grey) COVID-19 children.

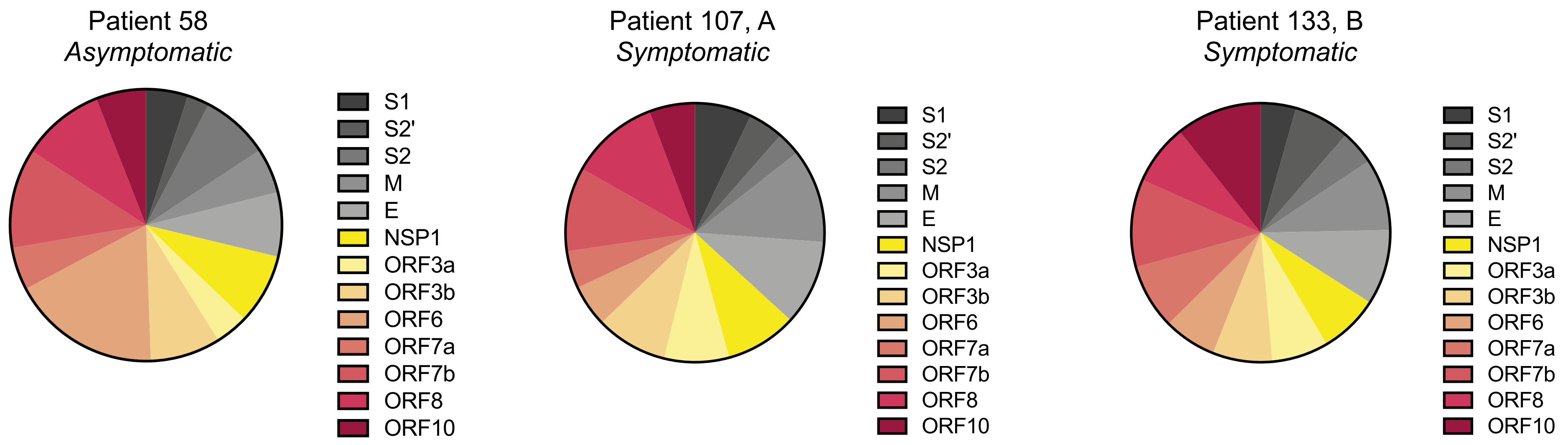


**Supplemental Figure 6.** Single pie charts of the cumulative antibody responses to the relevant SARS-CoV-2 structural and non-structural protein antigens (excluding N) for the COVID-19 pediatric patients producing the highest levels of IFN-α. (from Figure 7).

**Supplemental Table 1.** Correlation between variables and Principal Components 1 and 2 (Dimension 1 and 2 of 14, represented in Figure 3d-f).

| Dimension | Variable | Correlation | p.value |
|---|---|---|---|
| 1 | N | 0.5921686 | 0.00000 |
| 1 | S1 | 0.0906619 | 0.10884 |
| 1 | S2’ | 0.1981899 | 0.00041 |
| 1 | S2 | 0.4552057 | 0.00000 |
| 1 | ORF7a | 0.5495707 | 0.00000 |
| 1 | ORF3b | 0.6786038 | 0.00000 |
| 1 | ORF7b | -0.0681466 | 0.22853 |
| 1 | NSP1 | 0.6656403 | 0.00000 |
| 1 | ORF10 | -0.2925584 | 0.00000 |
| 1 | E | 0.275052 | 0.00000 |
| 1 | M | -0.1509175 | 0.00739 |
| 1 | ORF6 | -0.3774847 | 0.00000 |
| 1 | ORF8 | 0.6342754 | 0.00000 |
| 1 | ORF3a | 0.6259408 | 0.00000 |
| 2 | N | -0.0821349 | 0.14648 |
| 2 | S1 | 0.6981859 | 0.00000 |
| 2 | S2’ | 0.131899 | 0.01938 |
| 2 | S2 | 0.3052223 | 0.00000 |
| 2 | ORF7a | 0.5271245 | 0.00000 |
| 2 | ORF3b | 0.3465305 | 0.00000 |
| 2 | ORF7b | 0.5554244 | 0.00000 |
| 2 | NSP1 | -0.1296366 | 0.02158 |
| 2 | ORF10 | 0.608538 | 0.00000 |
| 2 | E | -0.44982 | 0.00000 |
| 2 | M | 0.3325727 | 0.00000 |
| 2 | ORF6 | 0.2425214 | 0.00001 |
| 2 | ORF8 | -0.1286999 | 0.02255 |
| 2 | ORF3a | -0.0883815 | 0.11807 |
